# Intrahousehold dynamics in South Asia: understanding the relationships among women’s empowerment, task sharing, women’s decision making, and diets

**DOI:** 10.1101/2025.01.08.25320196

**Authors:** Neha Kumar, Agnes Quisumbing, Swetha Manohar, Archis Banerjee, Shivani Gupta, Alka Chauhan, Sharvari Patwardhan, Uma Koirala, TAFSSA collaborators

**Author notes:** The Transforming Agri Food Systems in South Asia collaborators include: Avula, Rasmi (International Food Policy Research Institute); Banerjee, Anurag (International Water Management Institute); Banerjee, Archis (International Food Policy Research Institute); Chakrabarti, Suman (International Food Policy Research Institute); Chakraborty, Shreya (International Water Management Institute); Chellattan Veettil, Prakashan (International Rice Research Institute); Gupta, Ishika (International Rice Research Institute); Headey, Derek (International Food Policy Research Institute); Kabir, ANM Faijul (International Rice Research Institute); Kamal, Mustafa (International Maize and Wheat Improvement Center); Karki, Saral (International Maize and Wheat Improvement Center); Khan, Nur-A-Mahajabin (International Maize and Wheat Improvement Center); Kim, Sunny (International Food Policy Research Institute); Kishore, Avinash (International Food Policy Research Institute); Krupnik, Timothy Joseph (International Maize and Wheat Improvement Center); Laing, Alison (International Maize and Wheat Improvement Center); Menon, Purnima (International Food Policy Research Institute); Neupane, Sumanta (International Food Policy Research Institute); Tek Bahadur (International Maize and Wheat Improvement Center); Sarswat, Esha (International Food Policy Research Institute); Scott, Samuel (International Food Policy Research Institute); Sapkota Zainab, Afrin (International Rice Research Institute).

## Abstract

Despite the growing evidence on women’s roles in agriculture and nutrition, interlinkages between women’s empowerment, gendered task allocation, and nutrition are rarely studied together. Using data from the Transforming Agrifood Systems in South Asia (TAFSSA), a household survey that used a “plate to farm” assessment approach in three countries (Bangladesh, India and Nepal), the paper investigates the associations between women’s empowerment, gendered task allocation, women’s decision-making, and women’s diets. Our findings reveal complex and context-specific differences in associations between task allocation, decision-making and women’s empowerment. While agency in women’s decision-making is positively associated with empowerment in all three country contexts, associations between gendered task allocation and empowerment vary. The share of tasks performed by females, particularly in agriculture and food preparation) is positively associated with women’s empowerment, but the proportion of tasks shared equally between males and females does not necessarily empower women. Gendered task allocation and women’s empowerment are not significantly associated with women’s diets in the three countries, owing to the greater importance of broader socio-economic and context-specific factors such as wealth, education, and regional factors in explaining the variance in dietary outcomes. These findings highlight the need to take a holistic approach that addresses gender norms and household resource constraints to improving women’s empowerment, while also addressing local accessibility/availability of nutritious foods to enhance the quality of women’s diets.

## INTRODUCTION

A growing literature highlights the important linkages between women’s roles in agriculture and nutritional outcomes. For example, in the agriculture-nutrition framework developed by Ruel and Alderman (2013), three of the six identified pathways through which agricultural interventions can impact nutrition focus explicitly on women, notably women’s social status and empowerment through increased access to and control over resources; women’s time through participation in agriculture, which can be either positive or negative for their own nutrition and that of their children; and women’s health and nutrition through engagement in agriculture. These three pathways are interrelated. Women’s social status and empowerment can affect how much time she spends on various activities, while her allocation of time affects her empowerment status. Her decision-making power over different household domains could affect her time allocation as well as well-being outcomes such as her own diet and nutrition. Yet, these phenomena are rarely studied together owing to the lack of conceptual frameworks that simultaneously address these issues as well as the paucity of data on empowerment, time use, and nutrition on the same individuals. Moreover, where issues of women’s empowerment, time use, and nutrition are analyzed, linkages are usually drawn between women’s time use and empowerment and child nutrition, not women’s own nutritional status. This reflects the prevailing view that women’s empowerment is instrumental to child nutrition, not that it has intrinsic value forwomen’s own well-being. Yet, time use patterns are an important factor affecting food consumption practices and nutritional outcomes (Hull, 2013). Time spent in both productive and reproductive activities can contribute to time poverty among women (Blackden and Wodon, 2006; Seymour et al. 2020), decreasing the time available for production, procurement and preparation of nutritious food affecting women’s dietary intake (Johnston et al., 2018).

After the adoption of the women’s empowerment and gender equality as a Sustainable Development Goal (SDG5), the 2010s have seen rapid growth in the development of empowerment metrics (see Elias et al. 2021 for a review), many of which draw on Kabeer’s (1999) definition of empowerment as a process of making strategic life decisions, particularly for those who had previously been denied this ability.

Kabeer’s framework further defines empowerment as comprising agency, resources, and achievements. This framework provides a convenient point of departure for our analysis. Within this framework, time can be viewed as a resource, decisions regarding time allocation represent the exercise of agency, and dietary and nutrition outcomes are achievements attained through the deployment of resources and the exercise of agency.

Recently developed empowerment metrics conceptualize empowerment as a function of the amount of time spent on different types of activities (Alkire et al. 2013; Malapit et al. 2019; Eissler et al. 2022). For example, the Women’s Empowerment in Agriculture Index (WEAI) and project-level WEAI (pro-WEAI) include indicators related to an individual’s workload, which define someone as less empowered if they spend more than 10.5 hours/day on paid and unpaid work. However, purely quantitative approaches to measuring time use and its relationship with empowerment do not capture the subtleties in the decisions and tradeoffs women make around time use. Eissler et al. argue that automatically categorizing those with heavy time burdens as disempowered can mask key aspects of agency. They propose a new concept of “time-use agency,” defined as “the confidence and ability to make and act upon strategic choices about how to allocate one’s time.” Unpacking empowerment-time use relationships also requires a better understanding of how tasks are shared within households and whether task sharing contributes to or detracts from individual agency. However, time use surveys often collect information only on a subset of household members, such as the primary male and female respondent interviewed in the WEAI and its variants. As such, these do not provide information about task-sharing.

We take advantage of a recently completed survey of agricultural households in selected study sites in Bangladesh, India, and Nepal to analyze issues of empowerment, task allocation, decision-making, and women’s diets. The Transforming Agrifood Systems in South Asia (TAFSSA) district agrifood systems assessment was designed to provide a reliable, accessible, and integrated evidence base that links farm production, market access, dietary patterns, climate risk responses, and natural resource management with gender as a cross-cutting issue in rural areas of Bangladesh, India, and Nepal. Notably, the TAFSSA survey includes a task allocation module that obtains information on how different tasks are performed and shared within the household and how female respondents within these households perceive the notion of leisure, work pressure, and support with chores from other household members. Along with detailed data on domains of household decision-making and diets, this data set provides the opportunity to study the interlinkages among women’s empowerment, task allocation, decision-making, and diets. In this paper we examine some of these relationships to understand the linkages between women’s empowerment, their decision-making power and how tasks are performed and shared within the household. In addition, we examine if women’s empowerment and how tasks are shared within the household have any implications for women’s diets.

We describe the data and methods in section 2 and provide descriptive results in section 3. The regression results are presented in section 4 and section 5 discusses these findings. And, section 6 concludes.

## DATA

### The local agri-food system assessment survey

The data for this analysis comes from the Transforming Agrifood Systems in South Asia (TAFSSA) district agrifood systems assessment(Gupta et al. 2022). This assessment was designed to provide a reliable, accessible, and integrated evidence base that links farm production, market access, dietary patterns, climate risk responses, and natural resource management with gender as a cross-cutting issue in rural areas of Bangladesh, India, and Nepal. The survey was implemented in two districts in Bangladesh (Rajshahi and Rangpur), one in India (Nalanda), and two in Nepal (Banke and Surkhet) between February-May 2023. Data collection was carried out by Data Analysis and Technical Assistance in Bangladesh, Kabil Professional Services in India, and Institute for Integrated Development studies in Nepal. Survey sites were selected from TAFSSA learning locations facing significant poverty, malnutrition, social inequity, environmental degradation, and climate risks, with potential for substantial development impact (Gupta et al., 2022). Across all districts, 50 villages in Bangladesh and India, and 25 wards in Nepal, representing the primary sampling unit (PSU), were selected within each district with a probability proportional to the number of households in each village or ward. Within each village, a household listing was conducted to identify households with at least one adolescent (10–19 years old) member. From the list of households with adolescents, 20 households were randomly invited to participate in the survey.

Therefore, the data is representative of rural households from the included districts with at least one adolescent and are not representative of rural households in each country. The analysis presented in this paper should therefore not be interpreted as representing overall patterns for each country.

The agrifood systems assessment included a household survey with interviews conducted with three respondents in each household- one adult female aged 20+ years, one adult male aged 20+ years, and one adolescent aged 10-19 years. When multiple adolescents were living in a household, the oldest adolescent was selected. In some households, an adult male was not available (often due to migration for work). In such households, the female was the only adult respondent. After the purpose of the survey was explained to survey respondents, verbal informed consent was obtained by the enumerators from respondents. The survey included questions on demographic composition of the household, educational attainment and marital status of all household members, agricultural production, and dietary intake (described below). The primary adult female respondent was also asked how different tasks within the household were performed and shared between household members, decision-making, and a set of questions to assess their empowerment (described below). The task allocation module obtains information on how different tasks are performed and shared within the household (between themselves and/or other male and female adult and adolescent household members). Additionally,data was collected on how female respondents within these households perceive the notion of leisure, work pressure, and support with chores from other household members.

The TAFSSA survey was administered using computer-assisted personal interviewing (CAPI) techniques, and based on time stamps, the time needed to administer the task allocation module in the survey ranged from 3.88 minutes in India to 5.02 minutes in Nepal. Findings on each of the TAFSSA study sites are presented in Banerjee et al. (2023a, 2023b, 2023c, 2023d, 2023e); the analysis in this paper aggregates across sites within each country.

### Definition of key variables

We define the following variables that we analyze: (1) women’s empowerment; (2) task allocation; (3) decision-making; and (4) diets.

*Women’s Empowerment:* The women’s empowerment measure used is an index defined as a normalized sum of binary indicators across five domains: attitudes about domestic violence, financial independence, freedom of mobility, social support, and perceived work pressure. Each domain indicator takes the value of 1 if the primary respondent woman’s responses indicate the presence of the desired condition, or 0 otherwise.

1. Attitudes about domestic violence: Includes responses to attitude towards acceptance of domestic violence; the indicator takes on a value of 1 if she considers it unacceptable in any of the six situations listed, and 0 otherwise.
2. Financial independence: If a woman reports having the ability to use her money freely and has either a bank or mobile bank account, the indicator takes on a value of 1 and 0 otherwise.
3. Freedom of mobility: If a womant reports being able to travel to all listed locations without needing permission from her husband and/or another family member, the indicator takes on a value of 1, and 0 otherwise.
4. Family social support: If a woman talks to or sees her family members outside her home at least once a month, the indicator takes on a value of 1, and 0 otherwise.
5. Perceived work pressure: If the woman respondent perceives no work pressure or workload, the indicator takes on a value of 1, and 0 otherwise.

*Gendered Task Allocation:* To explore gendered task allocation and division of labor within households, we measure three indicators using three normalized scores. We use the biological distinction (males vs. females) because the category includes men and boys in the former, and women and girls in the latter.

1. <u>Male-performed tasks</u>: This variable is defined as the number of household tasks that are performed entirely by males in a household as a proportion of the total number of tasks performed.
2. <u>Female-performed tasks</u>: This variable is defined as the proportion of household tasks performed only by females in a household, relative to the total number of household tasks.
3. <u>Equal task sharing:</u> This variable represents the number of household tasks that are equally shared between men and/or boys and women and/or girls in a household as a proportion of the total number of tasks performed by the household.

To further examine gendered task allocation, we divide the three primary measures of task allocation into sub-categories of tasks within the household (the Appendix contains a detailed list of tasks included in each category):

a. Agricultural Tasks: This sub-component measures the proportion of agricultural tasks performed by males, females, or shared equally, relative to the total number of agricultural tasks in the household. These tasks include crop cultivation, livestock rearing and fish rearing activities.
b. Food Preparation Tasks: This sub-component measures the proportion of food preparation tasks performed by males, females, or shared equally, relative to the total number of food preparation tasks in the household. These tasks include purchasing food items, fetching water or fuelwood, cleaning food and preparing food.
c. Care and Maintenance Tasks: This sub-component measures the proportion of care and domestic maintenance tasks performed by males, females, or shared equally, relative to the total number of these tasks in the household. These tasks include house cleaning/maintenance, washing clothes, caring for children or the elderly.

*Women’s Decision Making:* The woman’s decision-making power index is the normalized sum of binary indicators across five decision making domains: decisions over her own health, child related matters, health expenditure, agriculture, and purchasing and preparing food. Each domain indicator takes the value of 1 if the woman makes the decision solely or jointly with others, or 0 otherwise

1. Decisions about own health: Includes decisions about a woman’s own health, the number of children a woman should have, and whether a woman uses a contraceptive method.
2. Decisions about child-related matters: Includes decisions on sending female and male children to school, and decisions regarding their marriages.
3. Decisions about health expenditure: Includes decisions related to expenditures on women’s, men’s, and children’s health.
4. Decisions about agriculture: Includes choices related to what to grow in the home/kitchen garden, what crops to cultivate in the field (excluding homestead), and the use of fertilizer.
5. Decisions about purchasing and preparing food: Includes choices about what foods to buy for the household and what foods to prepare on a day-to-day basis.

*Women’s Diets:* The global diet quality score (GDQS) is a population-based metric of diet quality that has been validated to predict nutrient inadequacy and non-communicable disease (NCD) risk outcomes globally (Moursi et al. 2021). The metric consists of 25 food groups (see, A1): 16 healthy food groups, 2 food groups that are unhealthy when consumed in excess, and 7 unhealthy food groups. For 24 of the food groups, quantity consumed is classified into: low, medium, and high. For high-fat dairy, quantity of consumption is classified into four categories: low, medium, high, and very high. For each respondent, points are assigned based on the levels of consumption. For the 16 healthy food groups, more points are given for higher intake. For the 7 unhealthy food groups, more points are given for lower intake. For the 2 food groups that are healthy when consumed in moderation, points are given as follows: for the red meat food group, the most points are given for moderate consumption (but not low or high consumption); for the high fat dairy food group, points are given for moderate and high consumption (but not low or very high consumption). In our analysis, we look at dietary quality using three different GDQS indicators:

1. GDQS Total: This is a summary measure of overall dietary quality and is evaluated by summing the points across all 25 GDQS food groups, with a possible range of 0 to 49. A higher score indicates a healthier diet.
2. GDQS+: This sub-metric measures the intake of nutrient-rich and diverse food groups. It is calculated by summing points across the 16 healthy GDQS food groups (possible range of 0 to 32).
3. GDQS–: This sub metric captures the consumption of unhealthy or excessive food groups. It is calculated by summing points across the 7 unhealthy GDQS food groups and the 2 GDQS food groups that are unhealthy when consumed in excessive amounts (possible range of 0 to 17).

The GDQS total score can be used to classify respondents based on their risk of nutrient inadequacy and diet-related noncommunicable diseases as follows: low (GDQS Total ≥23), moderate (≥15 and <23), or high (<15).

*Other Controls*: The analysis incorporates several control variables at the household, individual, and community levels to account for socio-economic, demographic, and infrastructural factors that may influence the relationships between gendered task allocation, women’s empowerment, and dietary outcomes. At the individual level, we include the primary female respondent characteristics such as age, years of education, and employment status. Household characteristics include the age, education and employment status of the household head, the proportion of dependents, the total landholding of the household and wealth quintiles as proxies for economic status, and the total number of tasks performed by the household. At the community level, the analysis accounts for access to financial institutions, the availability of general markets and smaller retail stores for daily necessities, educational infrastructure for children, the presence of cooperatives providing resources, and whether the nearest town is less than 20 km away.

## METHODS

To investigate the relationship between women’s empowerment, gendered task allocation, and dietary outcomes across three South Asian countries (Bangladesh, India, and Nepal), we estimate two models for each country. The first model explores the relationship between gendered task allocation and decision-making, and women’s empowerment, while the second model assesses the associations between gendered task allocation, women’s empowerment, and women’s dietary outcomes. We recognize that the cross-sectional nature of our data and the lack of suitable instruments to address endogeneity prevents us from attributing causality. Moreover, empowerment measures may also be affected by the same factors that influence nutritional outcomes. While we employed Ordinary Least Squares (OLS) regressions similar to other studies exploring relationships between women’s empowerment and dietary outcomes (Malapit et al. 2015b; Sraboni and Quisumbing, 2018; Holland and Rammohan, 2019; Quisumbing et al. 2021) to explore the associations between women’s empowerment, gendered task allocation, and dietary outcomes, we treat our estimated coefficients as indicative of associations rather than causation. To account for potential confounding influences, we control for individual, household, and community characteristics. We also incorporate block-level fixed effects to account for unobserved location-specific heterogeneity.

### Model 1: Women’s Empowerment and Gendered Task Allocation

In this model, we investigate the role of gendered task allocation and women’s decision-making power in shaping women’s empowerment. We estimate the following equation using OLS:

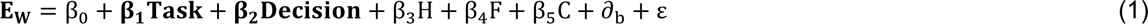

where E_W_ is the empowerment measure for the woman. Gendered task allocation, the key independent variable (represented by **TASK**) is measured using three normalized scores defined above: 1) Male-only tasks; (2) Female-only tasks; and (3) Equally-shared tasks. We include measures of women’s decision-making power (**Decision**) to proxy women’s agency in household decisions. We further include a set of control variables including primary respondent characteristics (**F**), household level characteristics (**H**) and community level characteristics (**C**). Finally, we include block fixed effects (∂_**b**_) to control for regional heterogeneity and cluster robust standard errors at the household level. β_1_, β_2_, β_3_, β_4_ and β_5_ are the parameters to be estimated; and ε is the error term.

### Model 2: Dietary Quality and Gendered Task Allocation

The second model explores the association between gendered task allocation, women’s empowerment, and women’s dietary outcomes. To evaluate this, we estimate the following equation:

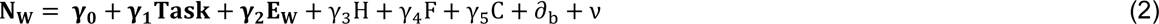

where **N**_W_ is a vector of woman-level diet outcomes measured using one of the three variants of the Global Diet Quality Score (GDQS) measures: 1) GDQS Total, 2) GDQS plus and GDQS minus. Gendered task allocation (**TASK**) measures and women’s empowerment (E_W_) are the main explanatory variables, as described in Model 1. We include household (**H**), female (**F**) and community-level (**C**) determinants as in model 1. Block fixed effects (∂_**b**_) control for regional differences across the three countries. γ_1_, γ_2_, γ_3_, γ_4_ and γ_5_ are the parameters to be estimated; and ν is the error term.

Our key coefficients of interest are **β**_1_and **β**_2_ from Model 1 (which captures how task allocation and women’s decision making are correlated with women’s empowerment outcomes), and **γ**_1_ and **γ**_2_ from Model 2 (which captures how task allocation and the women’s empowerment measure are correlated with women’s dietary outcomes). To examine the association between women’s empowerment, gendered task allocation, and dietary outcomes, we run separate regressions on each of the three task allocation measures in both models, to avoid collinearity among the different measures. Moreover, we also estimate separate parameters for each country to capture country-specific effects and account for heterogeneity in socio-cultural, economic, and institutional context.

To explore potential indirect effects of gendered task allocation and decision-making on dietary outcomes that work through women’s empowerment, we also conducted Structural Equation Modeling (SEM) to assess the simultaneous relationships between both the models. However, after finding no significant indirect effects, we estimate the models separately.

We also conducted Shapley-Owen decompositions to identify which predictors explained most variation in the outcome variables. This decomposition, calculated using the rego command in Stata version 18, decomposes the share of explained variance (measured by R-sq) into contributions by individual or groups of independent variables (Huettner and Sunder, 2012). Prior to conducting the regression analysis, we used the variance inflation factor (VIF) and correlation test (Pearson) to test the assumptions of linearity and multi-collinearity (Hair et al., 2010) between the control variables. We dropped variables which had VIF 10 or more before including the most important list of confounding variables. We also included only one of two variables if there was correlation of 0.7 or above between them. Owing to the possibility that the intrahousehold allocation of tasks may be affected by the same factors that influence the empowerment and nutritional outcomes, and our inability to find credible instruments for these indicators, we treat our estimated coefficients as suggestive of associations/correlations rather than causation.

## DESCRIPTIVE FINDINGS

In this section we present the summary statistics to characterize the sample. We use country names for brevity, but these statistics—and all the results in this paper—should be interpreted as findings from the specific study sites rather than representative of each country.

Table 1 presents the sample characteristics and covariates used in the analysis. Overall, the total numner of sampled study households included in the analyses was 1972, 978 and 1000 in Bangladesh, India and Nepal, respectively. The proportion of female-headed households differs significantly, ranging from 10.7% in Bangladesh to 27.2% in India and 46.2% in Nepal. In terms of the education levels among primary female respondents, we observe slight differences with the average number of years of education completed being 4.7 years in Bangladesh, 3.6 years in India, and 4.6 years in Nepal. Notably, Nepal records the highest employment rate for primary female respondents at 50.8%, followed by India at 38.9% and Bangladesh at 11.9%. Across the three countries, household sizes also vary with Bangladesh having the largest average household size (5.8). Household-level agricultural engagement remains high in all three sites. Eighty-two percent of households in India, 93.3% in Bangladesh, and 95.4% in Nepal reported some form of agricultural involvement. Infrastructure access vary between the three countries. Piped drinking water is accessible to 38.5% of Indian households, compared to just 6.1% in Nepal and 1.2% in Bangladesh. Similarly, cleaner cooking fuels such as LPG are commonly used in India (77.8%) and Nepal (76.3%). Reliance on traditional fuels like wood is more prevalent in Bangladesh, where only 23.8% of households report using LPG. Community-level resources further highlight the distinctions. For example, 40.9% of households in India report access to financial institutions, while the percentage of such households is 18% in Nepal and 10.1% in Bangladesh. Cooperatives are most widely accessible in Nepali communities (70%), followed by 63.2% in India and 52.4% in Bangladesh.

**Table 1.** Sample descriptives.

| Characteristics | Bangladesh | India | Nepal |
| --- | --- | --- | --- |
| **Sample Coverage** | Rajshahi, Rangpur | Nalanda | Surkhet, Banke |
| **Respondent Selection** | One adult female aged 20+ years, one adult male aged 20+ years, and one adolescent aged 10–19 years in each household. When multiple adolescents were living in a household, the oldest adolescent was selected. |  |  |
| **Number of Households in Estimation Sample** | 1,972 | 978 | 1,000 |
| **Individual-Level Characteristics** |  |  |  |
| Female-Headed Households (%) | 10.66 | 27.23 | 46.24 |
| Education of Household Head (years), mean (range) | 4.53 (0, 17.5) | 4.79 (0, 17.5) | 4.83 (0, 17.5) |
| Education of Primary Female Respondent (years), mean (range) | 4.66 (0, 17.5) | 3.61 (0, 17.5) | 4.62 (0, 17.5) |
| Employment Status of Household Head (%) | 92.99 | 89.04 | 67.1 |
| Employment Status of Primary Female Respondent (%) | 11.91 | 38.85 | 50.77 |
| **Household-Level Characteristics** |  |  |  |
| Average Household Size (Members) | 4.2 | 5.84 | 4.61 |
| Household Involved in Agriculture (%) | 93.26 | 82 | 95.4 |
| Household Age Dependency Ratio, mean (range) | 63.5 (0, 500) | 92.41 (0, 700) | 92.34 (0, 600) |
| Household's Source of Drinking Water (%) |  |  |  |
| - Tube well or borehole | 93.56 | 35.93 | 38.68 |
| - Piped into dwelling | 1.17 | 38.49 | 6.11 |
| Type of Fuel Used for Cooking in the Household (%) |  |  |  |
| - Wood | 90.57 | 60.29 | 93.69 |
| - Straw/Grass | 78.8 | 15.25 | 0.2 |
| - LPG/Natural Gas | 23.78 | 77.79 | 76.25 |
| Main Source of Income (%) |  |  |  |
| - Crop Cultivation | 39.15 | 38.65 | 25.9 |
| - Business | 24.9 | 9.92 | 12.2 |
| - Wages | 21.7 | 30.78 | 12.6 |
| Household Wealth Quintiles (%) |  |  |  |
| - First Quintile | 20.08 | 19.73 | 20 |
| - Second Quintile | 19.83 | 20.14 | 20 |
| - Third Quintile | 20.08 | 20.14 | 20 |
| - Fourth Quintile | 20.03 | 20.25 | 20 |
| - Fifth Quintile | 19.98 | 19.73 | 20 |
| Landholding Size (Acres), mean (range) | 0.57 (0, 16.67) | 0.48 (0, 10) | 0.64 (0, 12) |
| **Community-Level Characteristics** |  |  |  |
| Access to Financial Institutions (Bank in Community) (%) | 10.14 | 40.9 | 18 |
| Distance to Nearest Town (<20 km) (%) | 75.76 | 91.82 | 74 |
| Access to Educational Facilities (Middle School in Community) (%) | 22.31 | 85.69 | 76 |
| Availability of Markets/Bazaars (%) | 39.45 | 14.31 | 32 |
| Availability of Cooperatives (%) | 52.43 | 63.19 | 70 |

Notes: Authors calculations using the TAFSSA survey data

### Task sharing among household members

In all three countries and across different types of tasks, household members extensively share the undertaking of tasks (Banerjee et al. 2023a, 2023b, 2023c, 2023d, 2023e). For agricultural tasks, more task sharing between males and females is observed in Nepal as compared to India and Bangladesh. In Nepal, a higher proportion of households also reported agricultural tasks being undertaken solely by females, which is less likely in India and Bangladesh (except for tasks related to livestock rearing). Food preparation related tasks are performed primarily by females apart from tasks that entail leaving the homestead (for example, purchasing food or obtaining fuel for cooking). Care and maintenance tasks are also performed primarily by females. Tasks that required minor and/or major repairs are more likely to be undertaken by males and/or hired help.

Table 2 shows how tasks are performed by household members aggregated by task category by country. For example, the first cell in the column for Bangladesh shows that 19.34 percent of the agricultural tasks performed by households in Bangladesh are shared equally between males and females. Such sharing of tasks is much lower for food preparation (4.36%) and maintenance and care (7.79%) tasks, primarily undertaken by women, in Bangladesh. We observe relatively more sharing of tasks (across all categories) in India and Nepal as compared to Bangladesh. Nepal has the highest share of tasks being shared equally between males and females. We find that the largest proportion of food preparation related tasks are performed exclusively by females across all three countries (Bangladesh 77%, India 77% and Nepal 71%). Food preparation related tasks as also the tasks that are least likely to be performed exclusively by males across all three countries (Bangladesh 18.5%, India 9.5% and Nepal 4%). A significant proportion of maintenance and care tasks are also performed exclusively by females in all three countries (Bangladesh 69%, India 68% and Nepal 56%).

**Table 2.** Tasks performed by household members by type of tasks and country.

|  | Country |  |  | Total |
| --- | --- | --- | --- | --- |
|  | Bangladesh | India | Nepal |  |
|  | Mean Percentage |  |  |  |
| *Tasks performed by men only* |  |  |  |  |
| Agriculture | 39.10 | 26.12 | 6.57 | 27.75 |
| Food preparation | 18.54 | 9.48 | 4.42 | 12.74 |
| Maintenance & care | 20.13 | 12.66 | 9.70 | 15.65 |
| *Tasks performed by women only* |  |  |  |  |
| Agriculture | 38.49 | 34.69 | 56.39 | 42.31 |
| Food preparation | 77.05 | 77.15 | 71.37 | 75.65 |
| Maintenance & care | 69.01 | 64.08 | 56.32 | 64.60 |
| *Tasks performed by women and men* |  |  |  |  |
| Agriculture | 19.34 | 31.50 | 35.22 | 26.17 |
| Food preparation | 4.36 | 13.30 | 23.39 | 11.35 |
| Maintenance & care | 7.79 | 11.61 | 21.02 | 12.05 |

Notes: Authors calculations using the TAFSSA survey data

### Women’s decision making

We also examine women’s decision making in five domains: own health, child-related matters, health expenditures, agriculture and purchase/preparation of food. Figure 1 shows variation across countries in the proportion of households where women can make decisions either solely or jointly with other household members across the different domains. Over 90 percent of women in Bangladesh make decisions solely or jointly on all domains except for agriculture (where the corresponding percentage is 74%). In India, the pattern is similar to Bangladesh, but the proportion of sole and joint decision-making being much lower except for decisions related to food purchase and preparation). For example, 73-78% women reported being able to make decisions around own health, child related matters and health expenditures and only 36% reported making decisions on agriculture. Sole or joint decision-making among women in Nepal is not as high as in Bangladesh and India (except for decisions around agriculture where it is higher than both Bangladesh and India and decisions on own health where it is higher than India).

**Figure 1.**
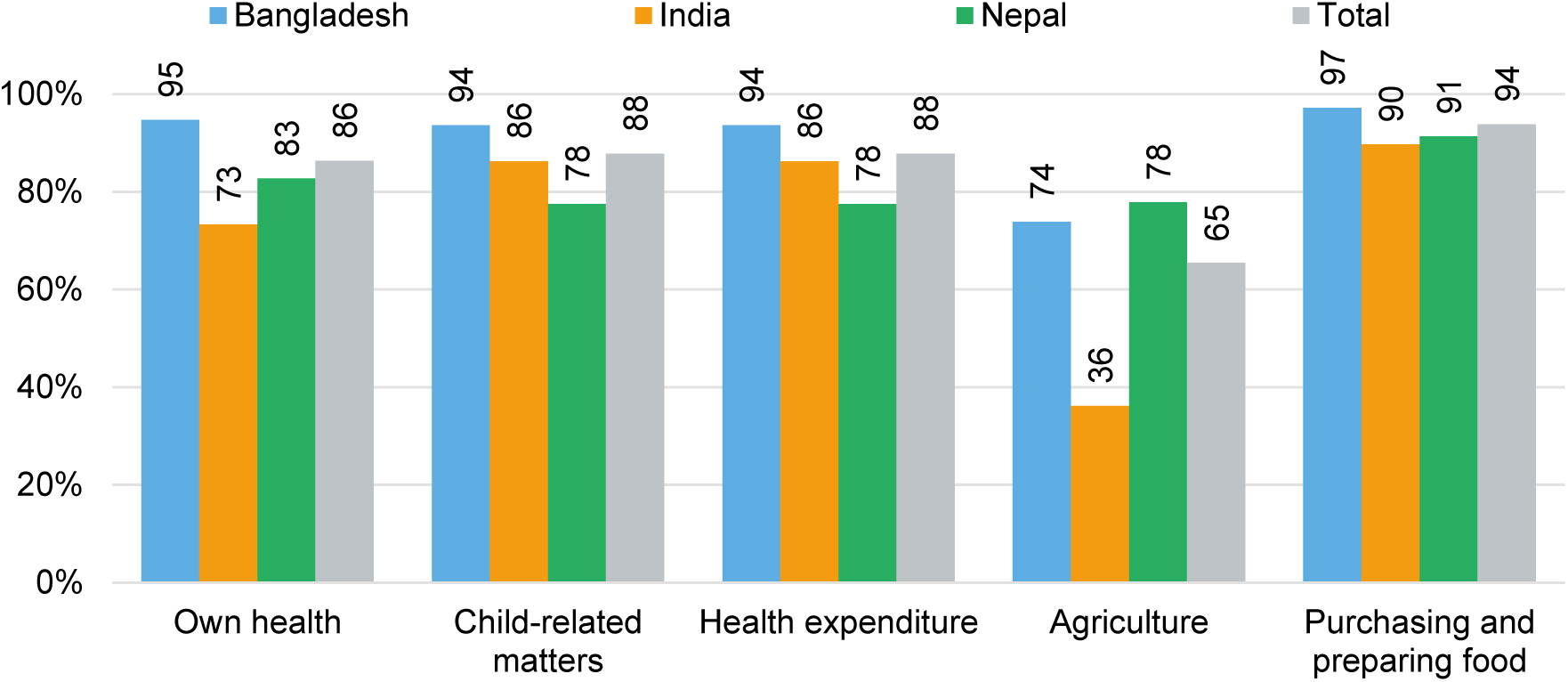
Distribution of households by women’s sole or joint decision-making across domains, by country

**Figure 1.**
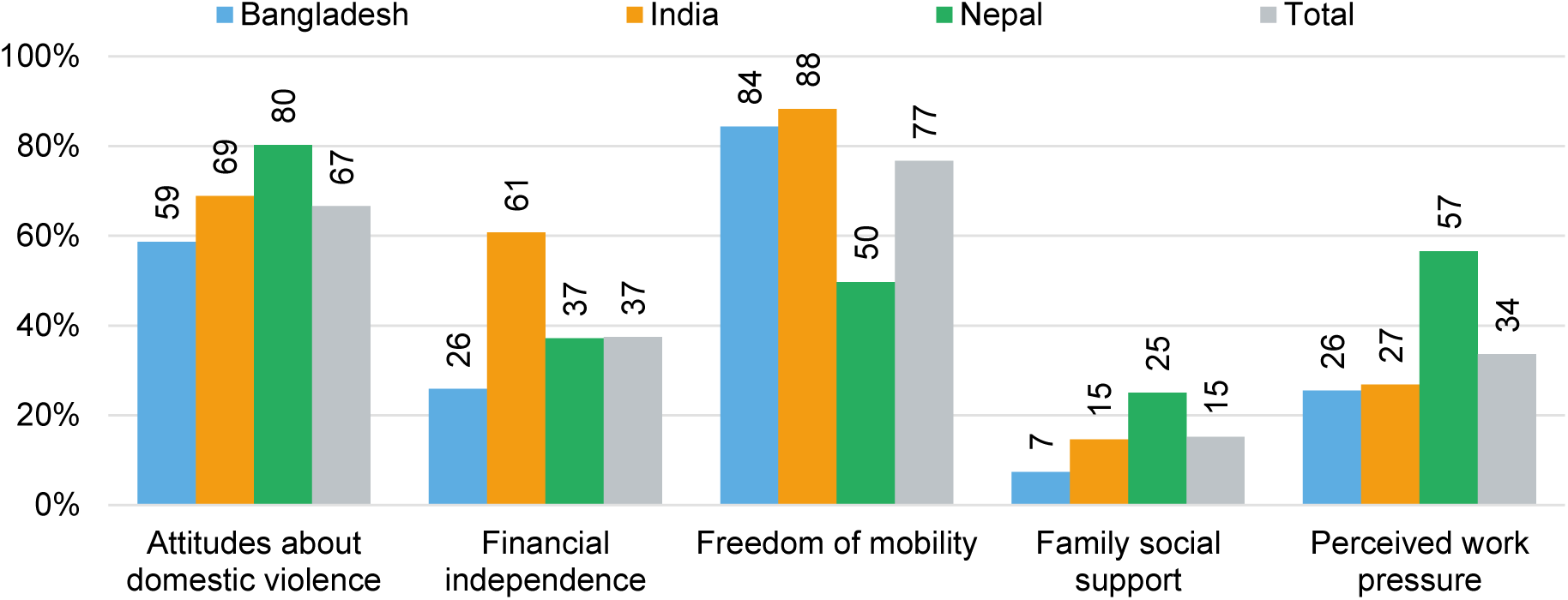
Distribution of women’s empowerment indicators by country

### Women’s empowerment

Figure 2 shows the distribution of the indicators underlying the women’s empowerment score. Overall, 67 percent of the women in the sample unequivocally reject any domestic violence (with the highest prevalence in Nepal at 80 percent and lowest in Bangladesh at 59 percent). Financial independence is highest among women in India with 61 percent reporting the ability to use their own money freely and having a bank account. Freedom of mobility is also highest in India with 80 percent of the women reporting that they don’t need permission to travel to various locations. Freedom of mobility is lowest among Nepali women where only 50 percent report not needing permission to travel. Overall, only 15 percent of women report having family social support. However, despite their relatively lower mobility Nepali women are more likely to report having family social support, with 25 percent saying that they spoke to or saw a family member in the past month. Additionally, Nepali women are also more likely to report no work pressure or excessive workload, unlike the corresponding proportion of women in India and Bangladesh, which is almost half of that in Nepal.

**Figure 2.**
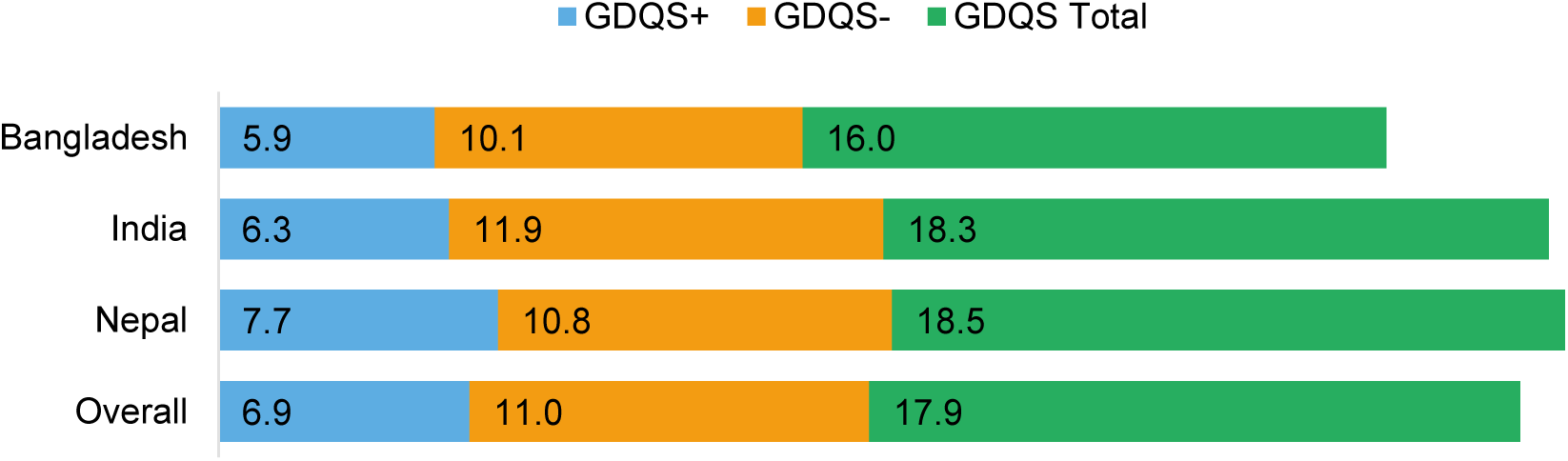
Mean GDQS scores (GDQS+, GDQS-, and GDQS Total) for primary female respondents across study countries and the overall sample

Figure 3 illustrates the mean GDQSscores: GDQS+, GDQS-, and GDQS Total, for the primary female respondents. Among the three countries, Nepal has the highest GDQS+ (healthy) score at 7.7, followed by India with 6.3 and Bangladesh with 5.9, with the overall GDQS+ average being 6.9. The GDQS- (unhealthy) scores are relatively close, ranging from 10.1 in Bangladesh to 11.9 in India, with an across country average of 11.0. For the GDQS Total, measuring overall diet quality, Nepal and India have a similar mean score of approximately 18, while Bangladesh have a lower duet quality of 16.0.

## REGRESSION RESULTS

### Women’s Empowerment and Gendered Task Allocation

Figures 4 to 6 present the estimated coefficients from regressions (equation 1) of the Women’s Empowerment Index on the three measures of gendered task allocation—male-only, female-only, and shared tasks—across Bangladesh, India, and Nepal. We also present disaggregated results from examining the effect of these intra-household task allocation categories on women’s empowerment in three domains: Agriculture, Food Preparation, and Care/Maintenance tasks. In each figure, we show the direction, magnitude and statistical significance of the associations.

**Figure 4.**
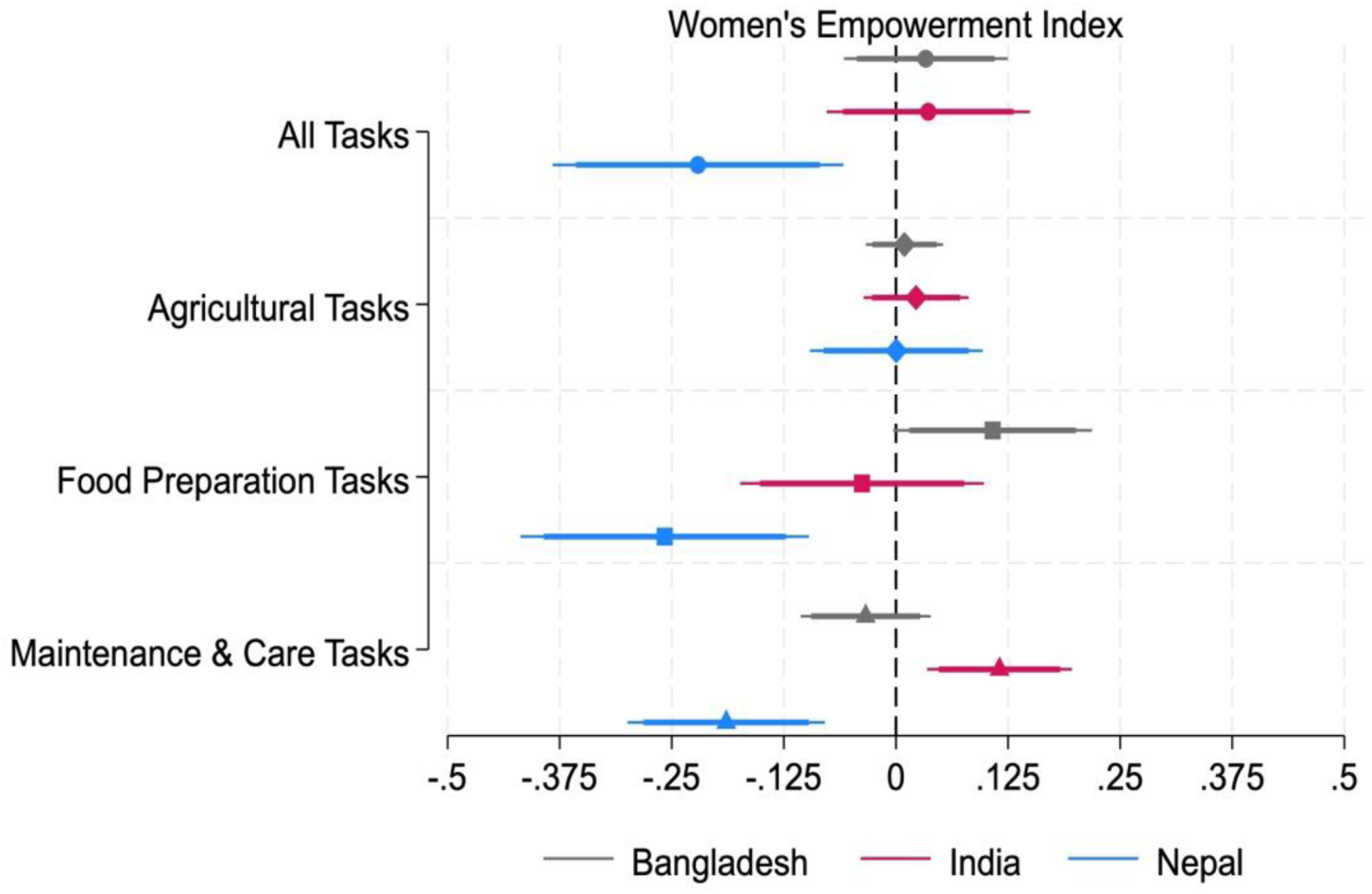
Coefficient estimates of the associations between women’s empowerment index and proportion of male-only tasks across task categories *Note: The figure reports estimated coefficients from regressions of the women’s empowerment index on the proportion of tasks performed only by males. 90% and 95% confidence intervals for all the estimates are represented by the narrow and wide lines, respectively*.

Figure 4, which presents coefficient estimates of the associations between the proportion of male-only tasks and the women’s empowerment index, reveals mixed findings across the three countries. Overall, we find a positive (although insignificant) association in Bangladesh and India, but a significant and negative relationship in Nepal. When we examine the different task categories, we find that the proportion of food preparation tasks performed only by males in Bangladesh and the proportion of care/maintenance tasks performed only by males in India are positively associated with women’s empowerment. In Nepal, however, the proportions of food preparation tasks and care/maintenance tasks performed only by males are negatively associated with women’s empowerment. The relationship between the proportion of agricultural tasks performed exclusively by males and women’s empowerment is insignificant.

Figure 5 shows that the relationship between the proportion of tasks performed exclusively by females and women’s empowerment is positive in all three countries. These positive associations are particularly strong in agricultural tasks in Bangladesh, food preparation tasks in India, and both agricultural and food preparation tasks in Nepal. We do not find any significant relationship between the proportion of care/maintenance tasks performed exclusively by females and their empowerment across the three countries.

**Figure 5.**
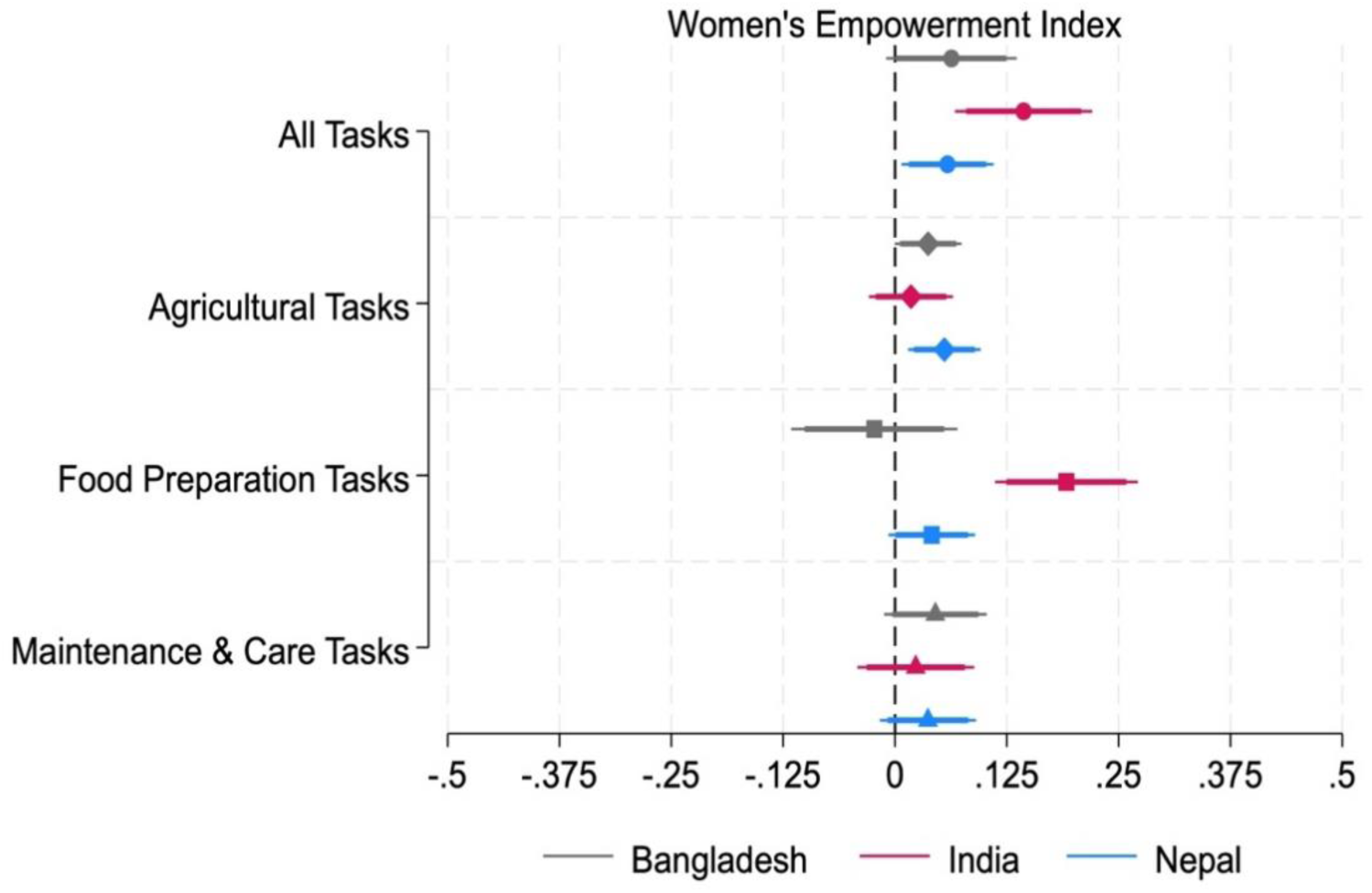
Coefficient estimates of the associations between women’s empowerment index and proportion of female-only tasks across task categories *Note: The figure reports estimated coefficients from regressions of the women’s empowerment index on the proportion of tasks performed only by females. 90% and 95% confidence intervals for all the estimates are represented by the narrow and wide lines, respectively*.

**Figure 6.**
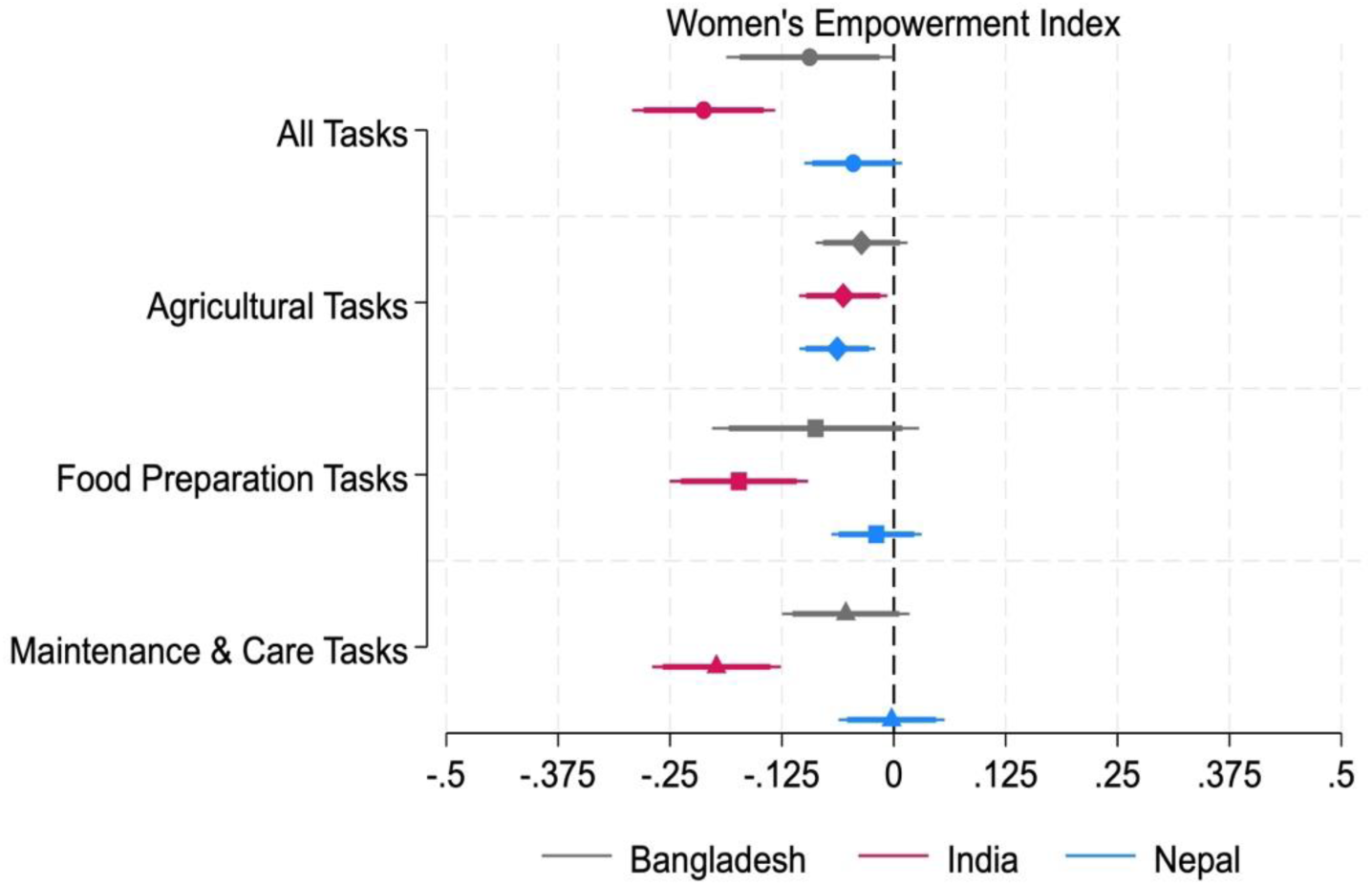
Coefficient estimates of the associations between women’s empowerment index and proportion of equally-shared tasks across task categories *Note: The figure reports estimated coefficients from regressions of the women’s empowerment index on the proportion of tasks equally-shared between males and females.90% and 95% confidence intervals for all the estimates are represented by the narrow and wide lines, respectively*.

Figure 6 relates the proportion of tasks that are shared equally between males and females and women’s empowerment. The proportion of equally shared tasks is significantly negatively associated with women’s empowerment in Bangladesh and India. In Nepal, however, while we note the same direction of association, thee finding is not statisticallysignificant. No significant associations were noted between women’s empowerment and the specific task categories of agriculture, food preparation, and care/maintenance.We calculate the Shapley/Owen values, which decomposes the explained variance (R-squared) of the model into individual/group contributions of the gendered task allocation and women’s decision-making variables along with other control variables (Huettner and Sunder, 2012). We report these only for the composite task indicators in Table A1-A3 in the Appendix. The overall R-squared values ranged from 0.13-0.19 across the different task indicators and country specifications, with the models explaining an acceptable portion of the variance in the empowerment outcomes.

The contribution of the male-only task allocation indicator to the overall variance in women’s empowerment (Table A1) varies across the three countries. In Nepal, it has a significant negative association (-0.221, *p*<0.01), contributing 6.86% to the model R-squared. In contrast, Bangladesh shows a positive association with a small contribution (0.033, 3.17% R-squared), while India shows a minimal contribution (0.036, 0.43% R-squared) to the variance in women’s empowerment, albeit with statistically insignificant effects. The proportion of tasks performed by females alone (Table A2) is positively and significantly associated with women’s empowerment with varying contributions across the three countries. In India, we find the strongest correlation (0.144, p<0.01), with female-only task allocation contributing 14.63% to the model’s explanatory power. Though smaller in magnitude, the associations in Nepal (0.059, *p*<0.05) and Bangladesh (0.063, p<0.10) still have meaningful contributions, accounting for 4.66 % and 6.45% of the explained variation in empowerment outcomes. The proportion of tasks that are equally shared (Table A3) has a consistently negative association with the women’s empowerment index across all three countries, although the strength of the association varies. In India, we find the strongest association with a coefficient of -0.212 (p<0.01), explaining 22.23% of the variation in empowerment.

However, in Bangladesh and Nepal, the proportion of tasks that are shared equally accounts for only a small proportion of the variation (2-3%) in women’s empowerment, although the association remains significant in Bangladesh (-0.094, p<0.05). Contributions of the women’s decision-making variable differ across specifications but range from 2.89 to 3.5% in Nepal, 3.1 to 3.5% in Bangladesh, and 7.3 to 8.3% in India.

The results from the Shapley decomposition analysis further reveals the importance of task allocation relative to other factors in explaining the variance in women’s empowerment differs across the three countries. Regional factors represented by the block fixed effects (combined) dominate the variation (30- 56%) in the empowerment indicator in all specifications across the three countries, indicating the influence of structural and community factors on empowerment outcomes.

In Bangladesh, the greatest contribution to women’s empowerment came from the total number of tasks that the household performs (11.5-13.1%), followed by the household head’s employment status (9- 10.5%), primary female’s education (7.6-7.9%) and women’s decision-making power (3%). The small to moderate contribution of task indicators relative to other factors suggests that socio-economic factors and the household’s overall task burden may be more important than the gendered allocation of tasks and women’s decision-making power. In contrast, in India, the gendered task allocation indicators contribute the most to the variance in women’s empowerment (0.4-22.2%), followed by decision-making power (7.3-8.3%), highest quintile of household wealth (3-4%), and household head’s education (2–3%). In Nepal, household wealth is the main contributor (22%), followed by primary female education (13-14%), female employment status (7-8%) and decision-making power (3–3.5%). This suggests that while task allocation matters for women’s empowerment, the associations are mediated by broader socio-economic inequalities.

### Dietary Quality and Gendered Task Allocation

Figures 7-9 present the estimated coefficients from regressions (equation 2) of the women’s diet indicators on the three measures of gendered task allocation—male-only, female-only, and shared tasks—and the women’s empowerment index across Bangladesh, India, and Nepal. We also present disaggregated results showing the corresponding coefficients of the intra-household task allocation categories (Agriculture, Food Preparation, and Care/Maintenance tasks) on women’s diets. Each figure shows the direction, magnitude and statistical significance of the associations. Since the overall patterns across the three variants of GQDS are similar, we present only the graphs for the GQDS total indicator.

**Figure 7.**
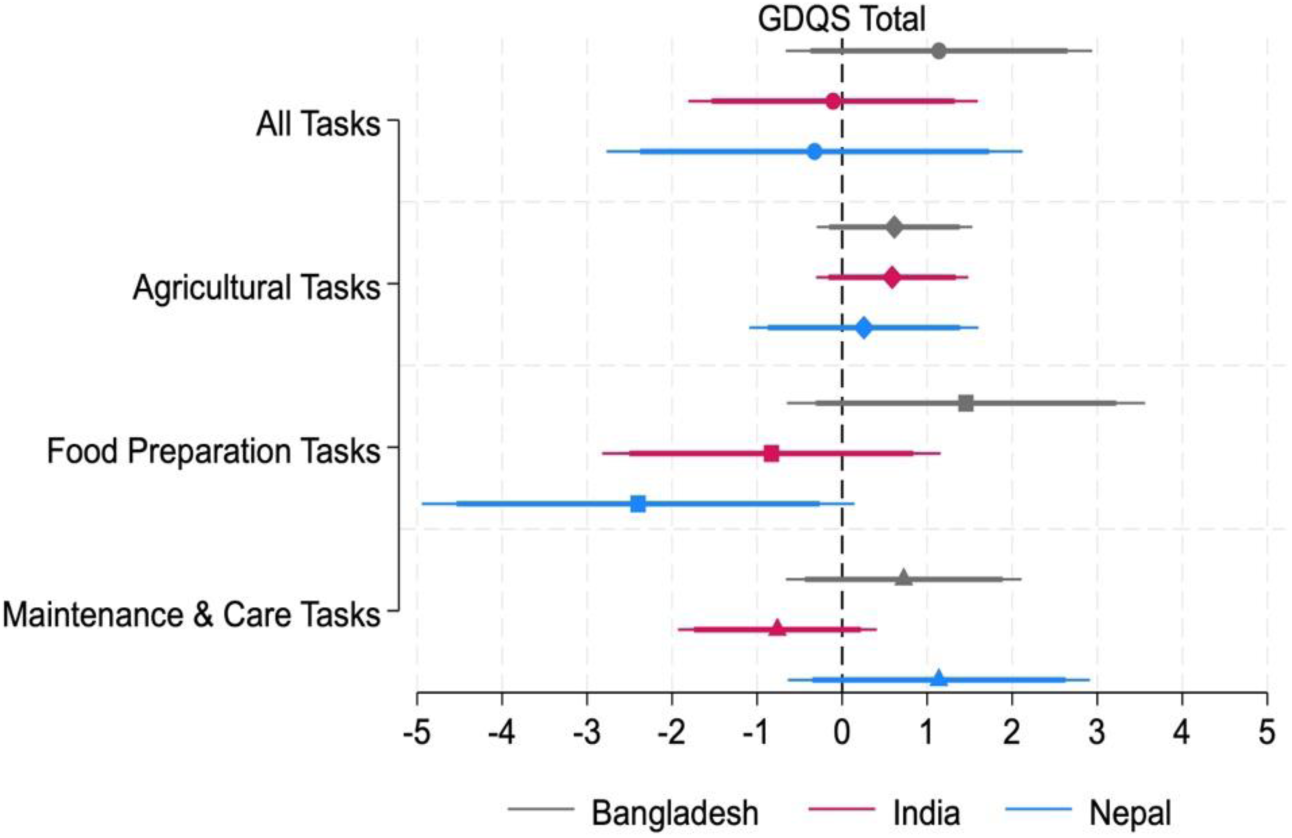
Coefficient estimates of the associations between women’s diets (GDQS Total) and proportion of male-only tasks across task categories *Note: The figure reports estimated coefficients from regressions of diet quality (GDQS Total) on the proportion of tasks performed only by males. 90% and 95% confidence intervals for all the estimates are represented by the narrow and wide lines, respectively*.

**Figure 8.**
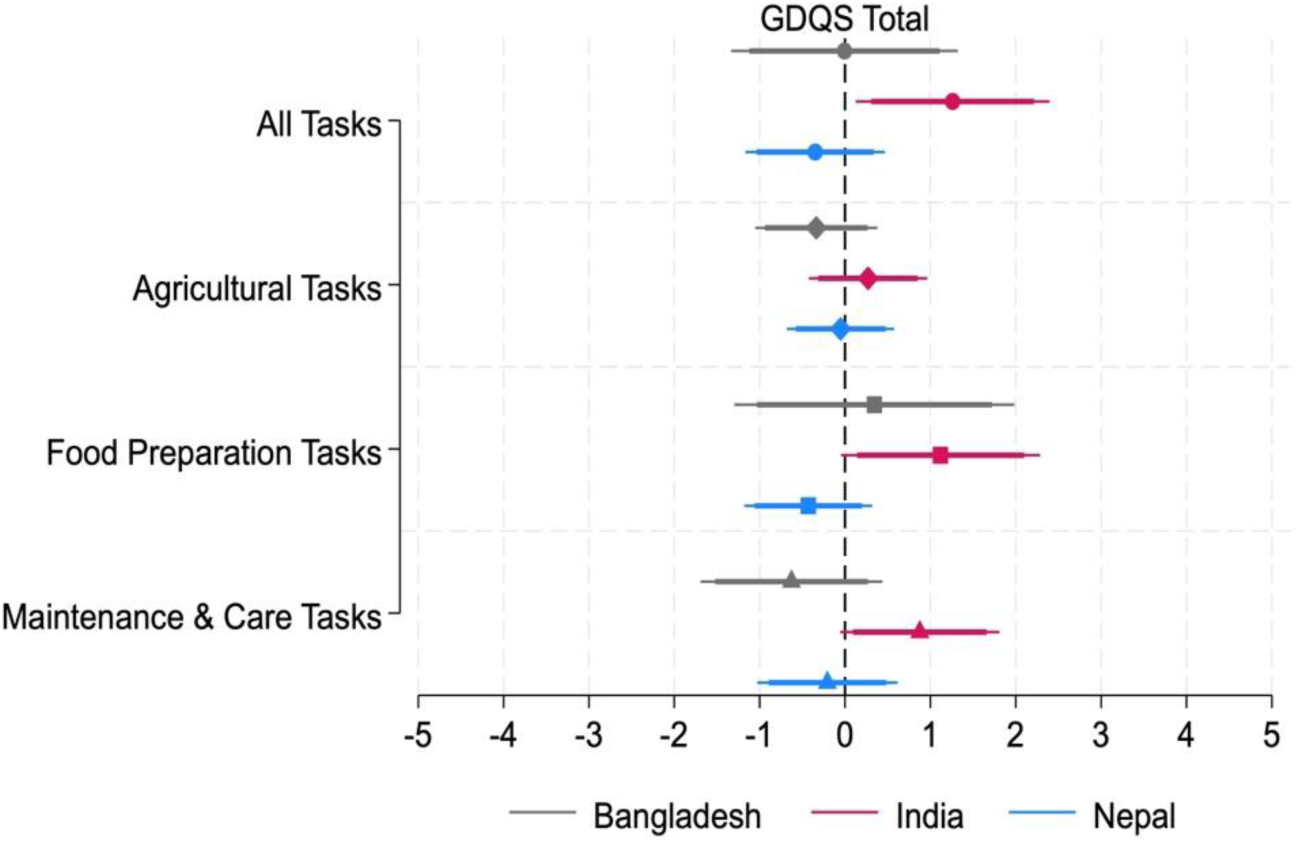
Coefficient estimates of the associations between women’s dietary diversity (GDQS Total) and proportion of female-only tasks across task categories *Note: The figure reports estimated coefficients from regressions of diet quality (GDQS Total) on proportion of tasks performed by only by females. 90% and 95% confidence intervals for all the estimates are represented by the narrow and wide lines, respectively*.

**Figure 9.**
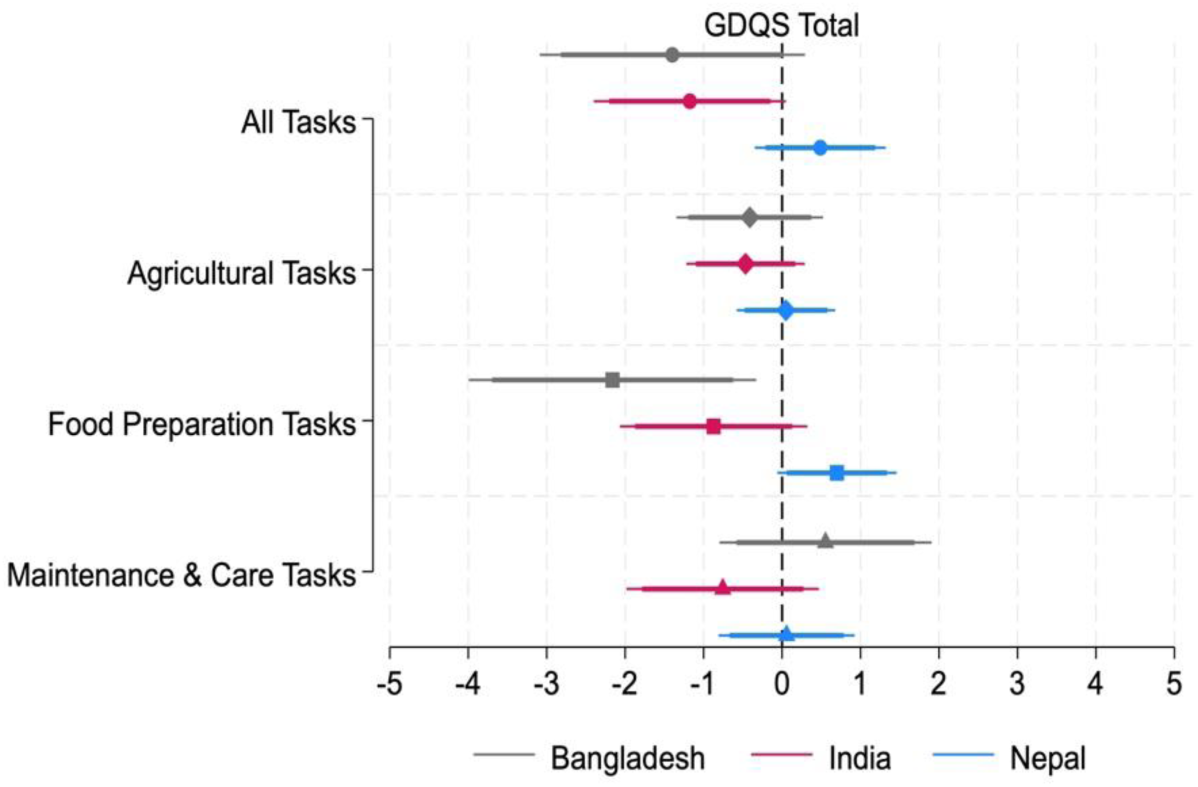
Coefficient estimates of the associations between women’s dietary diversity (GDQS Total) and proportion of equally-shared tasks across task categories *Note: The figure reports estimated coefficients from regressions of diet quality (GDQS Total) on the proportion of tasks equally shared between male and females. 90% and 95% confidence intervals for all the estimates are represented by the thin and thick lines, respectively*.

Overall, we do not observe any significant relationships between the proportion of tasks performed solely by males and women’s GDQS Total. In India, we find a positive (1.26, p<0.05) relationship between the proportion of tasks performed exclusively by females and women’s GDQS Total, primarily driven by the food preparation and maintenance and care tasks. We also find a negative (-1.17, p<0.1) relationship between the proportion of equally-shared tasks and women’s GDQS Total, in India. In Bangladesh, the equal-sharing task indicator has a negative relationship with women’s GDQS Total. Examining the different task domains, we find that this negative relationship is observed for equal task sharing in food preparation tasks only. In Nepal, we observe a positive relationship between equal task sharing in all tasks and GDQS Total (0.488) albeit the finding is not statistically significant.

The Shapley/Owen values for the diet models are shown in Tables A4-A6. As expected from the small magnitude of the estimated coefficients on the task allocation variables and their null impacts, task allocation variables account for a small proportion of the variation in women’s diets, regardless of the GQDS measure. The contribution of the proportion of male-only tasks ranges from 0.73 in Nepal to 2.99 in India, while the contribution of the proportion of female-only tasks ranges from 0.28 in Nepal to 2.05 in India. The proportion of tasks that are equally shared contributes only 0.5–3 percentage points to the variance in GDQS Total across Bangladesh, India, and Nepal. In contrast, in India, wealth explains over 45% of the variation in the GDQS Total whereas in Bangladesh and Nepal, 62 and 53 percent of the variation in GDQS Total, respectively, is explained by location fixed effects. Other covariates that explain the variation include the female respondent’s education (7% in Bangladesh, 8.8% in India and 9% in Nepal) and the household head’s education (4% in Bangladesh, 1.5% in India and 10% in Nepal).

## DISCUSSION

Our results highlight several important features of the relationships among women’s empowerment, decision-making, task allocation, and women’s diets.

### Women’s empowerment, task allocation, and decision-making

Regression results reveal complex relationships between women’s empowerment, task allocation, and decision-making. A consistent result is that the decision-making score, which reflects the scope of domains in which women decide solely or jointly with their spouses, has a positive association with women’s empowerment in all three countries. However, the association of gendered task allocation with women’s empowerment differs across country contexts. While the proportion of tasks performed by males only is positively associated with women’s empowerment in Bangladesh and India, in Nepal, this proportion is negatively associated with women’s empowerment. In all the countries, the proportion of tasks solely performed by females shows positive associations with women’s empowerment. Interestingly, the proportion of tasks shared equally between males and females has a negative relationship with women’s empowerment in Bangladesh and India.

The estimated relationships between the proportions of male-only, female-only, and shared tasks with women’s empowerment reveal that gender norms about task sharing are both context-specific and deeply entrenched. The positive association between the proportion of female-only tasks and women’s empowerment, particularly in contexts where they are responsible for agricultural or food-related tasks, reflects the beliefs that taking on traditional roles boosts perceived agency, even if these tasks may increase women’s workload. However, the negative association between the proportion of equally-shared tasks and women’s empowerment in India and Bangladesh runs counter to beliefs that task-sharing between men and women is more progressive and gender-equitable. It is possible that in countries where patriarchal norms are entrenched, traditional gender norms may constrain women’s ability to gain autonomy through task sharing. This relationship (between equally-shared tasks and women’s empowerment), however, is insignificant in Nepal, where gender norms may be modified by male outmigration. This suggests that while equitable sharing could be progressive in the longer run, societal norms are deeply entrenched, making it difficult to change behavior in the shorter run. For instance, perceptions of women’s empowerment could be tied to women taking ownership or control of performing these tasks in the household. Although it is widely perceived that both men and women have too little time for preferred activities and that excessive workloads are common sources of daily stress that may lead to negative consequences for individuals’ health and well-being (Almeida, 2005; Miedema et al., 2021), this view does not take into account their time use agency, or their confidence and ability to make and act upon strategic choices about how to allocate their time (Eissler et al. 2022). In this specific example, women-only tasks may convey more agency to women, who have a greater ability to control their time rather than if they had to share a task with men. A study of mothers-in-laws and daughters-in-law in Nepal (Doss et al. 2022) found that, although both do not work significantly different hours from each other, as revealed by quantitative time use data, daughters-in-law feel they are doing many more hours of work and, more importantly, feel they have little or no control over their work. In other words, they have less autonomy over work and thus less agency.

The Shapley decomposition results provide important insights in interpreting the findings on the association between gendered task indicators, women’s decision-making, and women’s empowerment. First, the contribution of task allocation to the variance in women’s empowerment is small in magnitude compared to other factors. The contribution of proportion of male-only tasks to empowerment ranges from 0.4% in India to 6.9% in Nepal. The contribution of female-only tasks ranges from 4.7% in Nepal to 14.6% in India. The contribution of the proportion of shared tasks is small in Nepal and Bangladesh, at 2.39% and 2.96%, respectively, but a large 22.23% in India. Second, while women’s decision-making has a positive association with women’s empowerment in all countries, its contribution to the variance is small, ranging from 3.1-3.5% in Bangladesh, 7.3-8.3% in India, and 2.9-3.5% in Nepal. Third, regional factors and factors associated with household wealth are more important than the gender-related variables in explaining the variance in empowerment outcomes. Indeed, regional factors dominate the variation (30- 56%) in the empowerment indicator in all specifications across the three countries, indicating the influence of structural and community factors on empowerment outcomes.

The results from the decomposition analysis reflect how broader socio-economic and structural factors can shape women’s empowerment and how the role of gender-based task allocation in influencing empowerment can vary depending on the social and cultural context of different countries. In India, where task-sharing has a negative impact, promoting gender-equitable task allocation alone may not improve empowerment and addressing women’s decision-making power and access to resources may be needed. In Bangladesh and Nepal, where structural factors like wealth and education dominate, addressing systemic barriers could better address the associations between gendered task allocation on women’s empowerment.

### Women’s diets, task allocation

Regression results show that task allocation and women’s empowerment variables do not significantly affect women’s diets in the three countries. This is consistent with the relatively small (in magnitude) contribution of these variables to the variation in women’s GQDS in all three countries, as reported above. For example, the contribution of the male-only task allocation variable ranges from 0.73% in Nepal, 2.5% in Bangladesh, and 2.96% in India. The corresponding numbers for the female-only tasks are 0.7% in Bangladesh, 2.05% in India, and 0.28% in Nepal. The proportion of shared tasks contributes only 1.97% in Bangladesh, 3.1% in India, and 0.45% in Nepal. Thus, it appears that even if the heavy workload of women is viewed as a deterrent to better diet quality, it does not appear to be the most important factor, at least in these study sites.

However, there are other factors that explain a larger share of the variation in diet quality among women in the three countries. For example, in India, wealth explains over 45% of the variation in the GDQS whereas in Bangladesh and Nepal, 62 and 53 percent of the variation in GDQS, respectively, is explained by regional factors. This suggests that factors associated with poverty, inequality and access may be more closely linked to poor dietary quality than task allocation and women’s empowerment.

These results are consistent with those obtained from a six-country study (Quisumbing et al. 2021) exploring the relationship between women’s empowerment and nutritional outcomes using the Women’s Empowerment in Agriculture Index (WEAI) (Alkire et al. 2013). Although the women’s diets and empowerment variables in the present study differ from those used in Quisumbing et al. (2021), WEAI contains indicators of time use and women’s decision-making as component indicators. Shapley decompositions also showed that that empowerment-related factors account for only a small portion of the variation in women’s diets, measured using the women’s dietary diversity score. In that study, which pools large data sets from six countries in South Asia and Africa, household wealth contributes between 65 to 67 percent of the variance in women’s dietary diversity in regressions using the empowerment score and the intrahousehold inequality score, followed by country fixed effects (around 16%), the number of women completing primary school in the household (around 8%), and whether the woman herself finished primary school (3.2 to 5.2%).

## CONCLUSION

Although the nature of the data limits causal inferences, our analysis provides valuable insights into the extent to which patterns of intrahousehold task sharing affect women’s empowerment and dietary outcomes.

The relationship between women’s empowerment and what women do within the household and how tasks are shared across members within the household is complex. When tasks are shared equally across genders, it is not necessarily empowering for women. Similarly, despite the concern about women’s workload, it is not obvious that a higher the share of tasks exclusively performed by women is disempowering. These relationships are context specific as seen above. However, agency in decision-making was linked positively with women’s empowerment in all three contexts. Regional factors (often capturing gender norms and differences in local conditions) explained a large share of variation in women’s empowerment. These findings, taken together, underscore the importance of understanding the local context first and accordingly addressing gender norms as a way forward in improving women’s empowerment.

The findings on women’s diets emphasize the importance of household wealth, regional factors (which may include differences in access to diverse and healthy foods aside from gender norms), and other socioeconomic factors such as educational attainment. To improve women’s diets, we need to take a holistic approach that addresses gender norms and women’s empowerment, household resource constraints, and local accessibility/availability of nutritious foods. While these results may disappoint those who think empowering women will solve all nutritional problems: despite its positive association with many outcomes, our study highlights the need to address underlying conditions of poverty and wealth inequality alongside efforts to empower women.

## Data Availability

All data produced are available online at https://www.cgiar.org/news-events/news/open-access-agrifood-system-data-from-4000-households-across-bangladesh-india-and-nepal/

## ACKNOWLEDGMENT

We acknowledge all funders who supported this research through their contributions to the CGIAR Trust Fund: www.cgiar.org/funders/. This research was supported by the Transforming Agri-Food Systems in South Asia initiative, under the Consultative Group on International Agricultural Research.

## APPENDIX

**A1.** Global Diet Quality Score food groups.

A1. Global Diet Quality Score food groups
| Healthy | Unhealthy in excessive amounts | Unhealthy |
| --- | --- | --- |
| 1. Citrus fruits<br>2. Deep orange fruits<br>3. Other fruits<br>4. Dark green leafy vegetables<br>5. Cruciferous vegetables<br>6. Deep orange vegetables<br>7. Other vegetables<br>8. Legumes<br>9. Deep orange tubers<br>10. Nuts and seeds<br>11. Whole grains<br>12. Liquid oils<br>13. Fish and shellfish<br>14. Poultry and game meat<br>15. Low-fat dairy<br>16. Eggs | 1. High-fat dairy<br>2. Red meat | 1. Processed meat<br>2. Refined grains and baked goods<br>3. Sweets and ice cream<br>4. Sugar-sweetened beverages<br>5. Juice<br>6. White roots and tubers<br>7. Purchased deep fried foods |

**A1.** List of different tasks in three main sub-categories.

A2. List of different tasks in three main sub-categories:
| Agricultural Tasks | Food Preparation Tasks | Care and Maintenance tasks |
| --- | --- | --- |
| <u>Crop cultivation tasks:</u><br>i) Preparing land<br>ii) Sowing/seeding<br>iii) Weeding<br>iv) Irrigation<br>v) Harvesting food<br>vi) Post-harvest processing (threshing, grinding, drying, fermenting, etc.)<br>vii) Post-harvest storage<br>viii) Taking the produce to the market to sell<br><u>Livestock rearing tasks:</u><br>ix) Caring for large livestock like cows and buffalo<br>x) Grazing small livestock like goats | i) Purchasing perishable food (fruits, vegetables, meat, etc.)<br>ii) Purchasing non-perishable food (grains, lentils, oil, spices, etc.)<br>iii) Storing food at home<br>iv) Fetching water for drinking or food preparation<br>v) Cleaning food before cooking<br>vi) Obtaining fuel wood for cooking<br>vii) Obtaining other fuel for cooking<br>viii) Making tea<br>ix) Cooking food | i) Cleaning floors<br>ii) Washing clothes<br>iii) Minor repairs to the house<br>iv) Major repairs to the house<br>v) Caring for elderly<br>vi) Care for ill household members<br>vii) Feeding young children<br>viii) Bathing children<br>ix) Playing with children<br>x) Taking children to school<br>xi) Watching children at home<br>xii) Helping children with schoolwork |
| xi) Washing/cleaning livestock<br>xii) Cleaning livestock shed/space<br>xiii) Handling livestock dung<br>(includes making dung cakes)<br>xiv) Gathering food for livestock<br>xv) Cooking food for livestock<br>xvi) Caring for poultry<br>xvii) Taking livestock products to the<br>market to sell<br><u>Fish rearing tasks:</u><br>xviii) Caring for small fishponds<br>xix) Caring for large fishponds | x) Preparing food for<br>children under 2<br>xi) Serving food<br>xii) Dealing with leftovers<br>xiii) Dish washing |  |

**Table A 1.** Women’s empowerment, women’s decision making, and the proportion of male-only tasks.

| Variables | Bangladesh |  |  | India |  |  | Nepal |  |  |
| --- | --- | --- | --- | --- | --- | --- | --- | --- | --- |
|  | Coeff. |  | Contribution to R2 (%)<sup>a</sup> | Coeff. |  | Contribution to R2 (%)<sup>a</sup> | Coeff. |  | Contribution to R2 (%)<sup>a</sup> |
| Male-only task score | 0.033 |  | 3.169 | 0.036 |  | 0.430 | -0.221 | *** | 6.857 |
| Women's decision-making power | 0.060 | ** | 3.131 | 0.081 | *** | 8.096 | 0.046 | * | 2.893 |
| Female Respondent's age | 0.001 |  | 0.433 | 0.000 |  | 0.988 | 0.000 |  | 0.927 |
| Female Respondent's education (years) | 0.005 | *** | 7.887 | 0.002 |  | 1.508 | 0.007 | *** | 12.857 |
| Female Respondent employed | 0.024 |  | 1.182 | 0.012 |  | 2.007 | 0.055 | *** | 7.277 |
| Head's age | -0.001 | * | 2.133 | -0.001 |  | 3.588 | 0.000 |  | 0.444 |
| Head's education (years) | 0.001 |  | 2.769 | -0.003 | ** | 3.121 | -0.001 |  | 3.035 |
| Head employed | -0.084 | *** | 10.146 | -0.037 |  | 2.553 | 0.006 |  | 1.084 |
| Age dependency ratio | 0.000 |  | 0.364 | 0.000 |  | 0.379 | 0.000 |  | 0.221 |
| Total land (acre) | -0.004 |  | 0.662 | 0.006 |  | 1.323 | 0.007 |  | 0.447 |
| Wealth quintile: 1 | -0.027 |  | 0.696 | -0.056 | ** | 3.944 | -0.145 | *** | 10.541 |
| Wealth quintile: 2 | -0.040 | ** | 2.770 | -0.029 |  | 0.465 | -0.131 | *** | 9.864 |
| Wealth quintile: 3 | -0.009 |  | 0.176 | -0.026 |  | 0.436 | -0.077 | *** | 1.810 |
| Wealth quintile: 4 | 0.001 |  | 0.557 | -0.018 |  | 0.254 | -0.028 |  | 2.166 |
| Total household tasks | -0.004 | *** | 13.097 | -0.001 |  | 0.695 | -0.002 |  | 0.822 |
| Bank branch in community | -0.033 | * | 1.135 | -0.007 |  | 0.436 | -0.009 |  | 2.985 |
| Market/bazaar in community | -0.018 |  | 0.379 | 0.003 |  | 0.361 | -0.006 |  | 0.764 |
| Kirana market in community | 0.011 |  | 0.821 | -0.017 |  | 9.282 | 0.033 |  | 3.504 |
| Middle school in community | 0.038 | ** | 1.258 | 0.017 |  | 1.083 | 0.065 |  | 0.324 |
| Cooperative in community | -0.007 |  | 0.482 | -0.010 |  | 1.623 | -0.071 |  | 0.640 |
| Nearest town < 20 km | -0.023 | * | 0.479 | -0.009 |  | 0.942 | -0.050 |  | 1.479 |
| Block fixed effects | n.a. |  | 46.273 | n.a. |  | 56.486 | n.a. |  | 29.061 |
| N | 1624 |  |  | 969 |  |  | 970 |  |  |

\*\*\* $p < .01$ , \*\* $p < .05$ , \* $p < .1$
<sup>a</sup>Shapley-Owen decomposition of contribution to R-squared. Grouped R-squareds presented for the block dummies.

**Table A 2.**
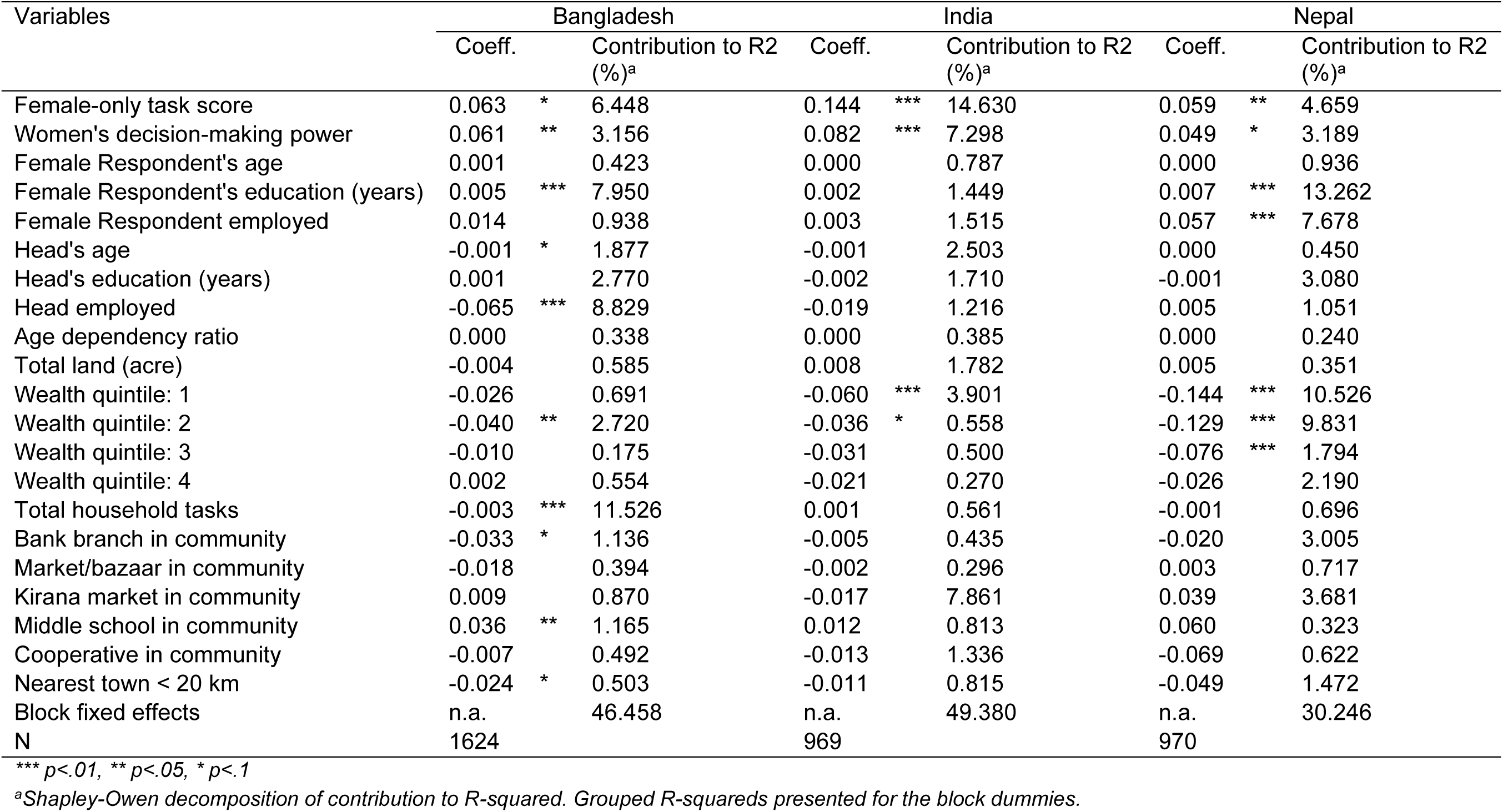
Women’s empowerment, women’s decision-making, and the proportion of female-only tasks.

**Table A 3.**
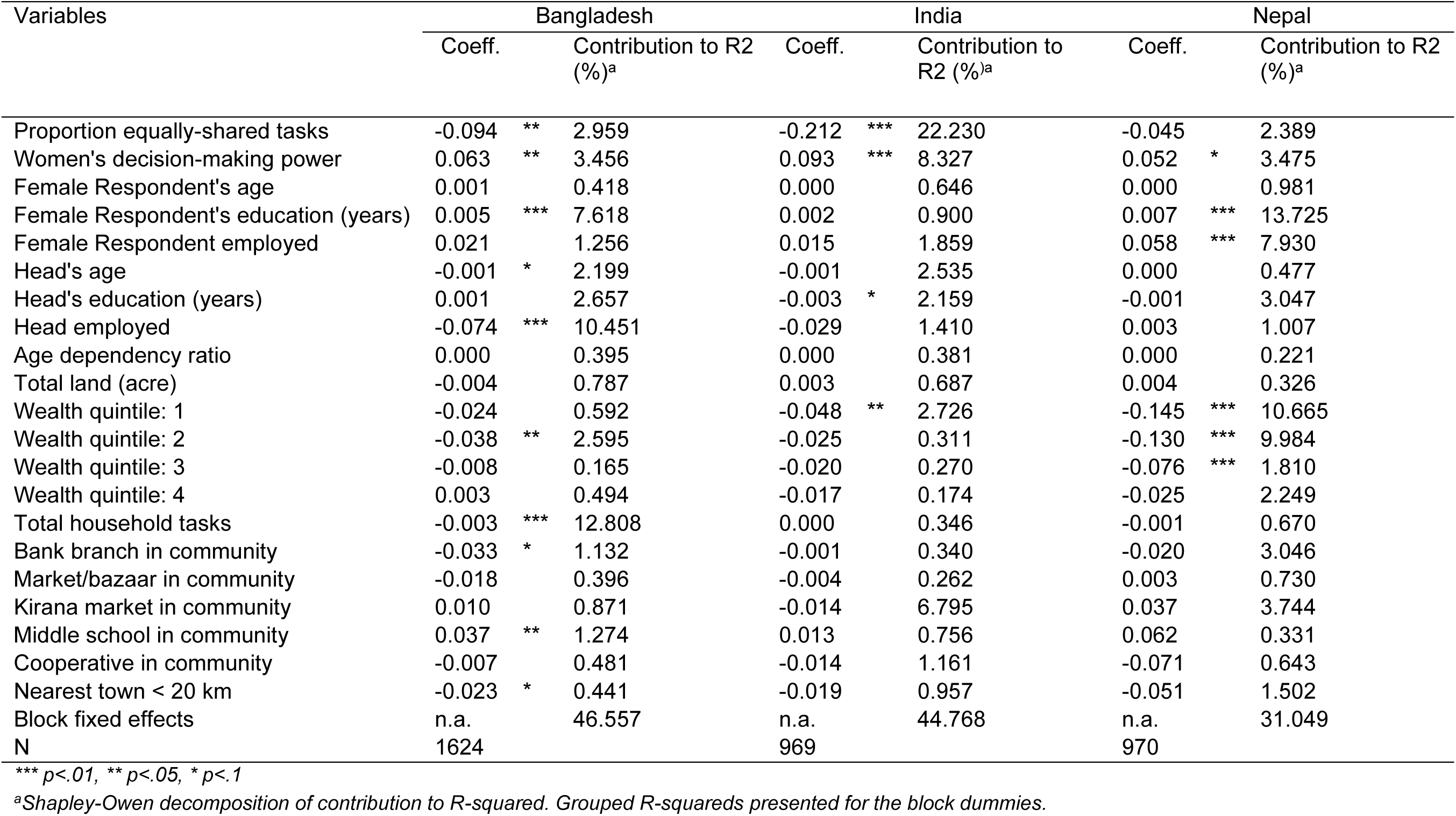
Women’s empowerment, women’s decision-making, and proportion of equally shared tasks.

**Table A 4.** Women’s diets (GDQS Total), women’s empowerment, and proportion of male-only tasks.

| Variables | Bangladesh |  | India |  | Nepal |  |
| --- | --- | --- | --- | --- | --- | --- |
|  | Coeff. | Contribution to R2 (%)<sup>a</sup> | Coeff. | Contribution to R2 (%)<sup>a</sup> | Coeff. | Contribution to R2 (%)<sup>a</sup> |
| Male-only task score | 1.138 | 2.450 | -0.107 | 2.958 | -0.325 | 0.733 |
| Women's empowerment score | 0.730 | 4.287 | 0.814 * | 2.647 | 0.078 | 0.331 |
| Female Respondent's age | 0.007 | 1.310 | -0.020 * | 2.611 | 0.030 ** | 4.388 |
| Female Respondent's education (years) | 0.048 * | 7.213 | 0.025 | 8.718 | 0.057 | 8.959 |
| Female Respondent employed | 0.356 | 1.609 | -0.297 | 4.101 | -0.585 ** | 2.075 |
| Head's age | 0.012 | 1.541 | 0.003 | 0.538 | 0.018 | 2.295 |
| Head's education (years) | 0.027 | 4.096 | -0.010 | 1.400 | 0.089 ** | 10.474 |
| Head employed | 0.482 | 2.220 | -0.229 | 0.738 | -0.094 | 0.669 |
| Age dependency ratio | -0.001 | 0.325 | -0.002 ** | 5.775 | 0.001 | 1.372 |
| Total land (acre) | -0.070 | 0.409 | 0.051 | 2.868 | -0.052 | 0.204 |
| Wealth quintile: 1 | -0.773 ** | 4.541 | -1.568 *** | 14.493 | -0.388 | 0.759 |
| Wealth quintile: 2 | -0.202 | 0.737 | -1.584 *** | 14.175 | -0.769 ** | 2.739 |
| Wealth quintile: 3 | -0.322 | 0.504 | -1.342 *** | 9.063 | -0.958 ** | 4.742 |
| Wealth quintile: 4 | -0.404 | 1.708 | -0.009 | 10.764 | -0.554 | 0.873 |
| Total household tasks | 0.011 | 1.127 | 0.015 | 1.619 | -0.001 | 0.127 |
| Bank branch in community | 0.061 | 0.184 | 0.269 | 0.354 | 0.819 | 0.900 |
| Market/bazaar in community | -0.022 | 0.431 | -0.143 | 0.365 | -0.264 | 0.699 |
| Kirana market in community | 0.172 | 1.452 | -0.445 | 0.692 | 0.201 | 0.780 |
| Middle school in community | 0.040 | 0.070 | -0.042 | 0.143 | -0.767 | 0.878 |
| Cooperative in community | 0.049 | 0.091 | -0.042 | 0.134 | -0.568 | 0.420 |
| Nearest town < 20 km | -0.036 | 1.596 | -0.098 | 0.076 | -0.558 | 1.925 |
| Block fixed effects | n.a. | 62.100 | n.a. | 15.765 | n.a. | 53.659 |
| N | 1624 |  | 969 |  | 970 |  |

\*\*\* $p < .01$ , \*\* $p < .05$ , \* $p < .1$
<sup>a</sup>Shapley-Owen decomposition of contribution to R-squared. Grouped R-squareds presented for the block dummies.

**Table A 5.** Women’s diets (GDQS Total), women’s empowerment, and proportion of female-only tasks.

| Variables | Bangladesh |  | India |  | Nepal |  |
| --- | --- | --- | --- | --- | --- | --- |
|  | Coeff. | Contribution to R2 (%)<sup>a</sup> | Coeff. | Contribution to R2 (%)<sup>a</sup> | Coeff. | Contribution to R2 (%)<sup>a</sup> |
| Female-only task score | -0.006 | 0.710 | 1.260 ** | 2.045 | -0.349 | 0.284 |
| Women's empowerment score | 0.741 | 4.330 | 0.687 | 2.433 | 0.122 | 0.380 |
| Female Respondent's age | 0.007 | 1.337 | -0.019 * | 2.554 | 0.031 ** | 4.389 |
| Female Respondent's education (years) | 0.048 | 7.362 | 0.022 | 8.777 | 0.062 | 9.101 |
| Female Respondent employed | 0.233 | 1.216 | -0.341 | 4.614 | -0.561 * | 1.975 |
| Head's age | 0.013 | 1.730 | 0.005 | 0.713 | 0.016 | 2.121 |
| Head's education (years) | 0.028 | 4.172 | 0.000 | 1.680 | 0.082 ** | 10.201 |
| Head employed | 0.704 * | 3.140 | -0.115 | 0.560 | -0.144 | 0.735 |
| Age dependency ratio | -0.001 | 0.498 | -0.003 *** | 6.135 | 0.002 | 1.521 |
| Total land (acre) | -0.059 | 0.440 | 0.056 | 3.213 | -0.063 | 0.236 |
| Wealth quintile: 1 | -0.775 ** | 4.759 | -1.601 *** | 14.344 | -0.403 | 0.759 |
| Wealth quintile: 2 | -0.204 | 0.733 | -1.633 *** | 14.030 | -0.787 ** | 2.751 |
| Wealth quintile: 3 | -0.322 | 0.512 | -1.379 *** | 9.066 | -0.965 ** | 4.718 |
| Wealth quintile: 4 | -0.388 | 1.664 | -0.027 | 10.438 | -0.544 | 0.846 |
| Total household tasks | 0.016 | 1.417 | 0.027 | 2.246 | -0.001 | 0.114 |
| Bank branch in community | 0.058 | 0.181 | 0.297 | 0.364 | 0.832 | 0.905 |
| Market/bazaar in community | -0.005 | 0.434 | -0.192 | 0.361 | -0.276 | 0.726 |
| Kirana market in community | 0.152 | 1.451 | -0.446 | 0.659 | 0.159 | 0.783 |
| Middle school in community | 0.020 | 0.064 | -0.080 | 0.125 | -0.725 | 0.857 |
| Cooperative in community | 0.041 | 0.084 | -0.069 | 0.128 | -0.580 | 0.423 |
| Nearest town < 20 km | -0.053 | 1.555 | -0.122 | 0.066 | -0.576 | 1.942 |
| Block fixed effects | n.a. | 62.210 | n.a. | 15.447 | n.a. | 54.233 |
| N | 1624 |  | 969 |  | 970 |  |

\*\*\* $p < .01$ , \*\* $p < .05$ , \* $p < .1$
<sup>a</sup>Shapley-Owen decomposition of contribution to R-squared. Grouped R-squareds presented for the block dummies.

**Table A 6.** Women’s diets (GDQS Total), women’s empowerment, and proportion of equally-shared tasks.

| Variables | Bangladesh |  | India |  | Nepal |  |
| --- | --- | --- | --- | --- | --- | --- |
|  | Coeff. | Contribution to R2 (%)<sup>a</sup> | Coeff. | Contribution to R2 (%)<sup>a</sup> | Coeff. | Contribution to R2 (%)<sup>a</sup> |
| Proportion equally-shared tasks | -1.399 | 1.969 | -1.175 * | 3.102 | 0.488 | 0.446 |
| Women's empowerment score | 0.705 | 3.955 | 0.664 | 2.082 | 0.122 | 0.394 |
| Female Respondent's age | 0.006 | 1.313 | -0.019 | 2.408 | 0.031 ** | 4.378 |
| Female Respondent's education (years) | 0.048 | 7.240 | 0.023 | 8.832 | 0.061 | 9.085 |
| Female Respondent employed | 0.233 | 1.128 | -0.250 | 4.245 | -0.563 * | 1.973 |
| Head's age | 0.014 | 1.870 | 0.003 | 0.717 | 0.016 | 2.112 |
| Head's education (years) | 0.028 | 4.023 | -0.008 | 1.655 | 0.083 ** | 10.133 |
| Head employed | 0.762 ** | 3.874 | -0.223 | 0.583 | -0.150 | 0.749 |
| Age dependency ratio | -0.001 | 0.705 | -0.003 ** | 6.429 | 0.002 | 1.567 |
| Total land (acre) | -0.064 | 0.471 | 0.029 | 2.951 | -0.058 | 0.241 |
| Wealth quintile: 1 | -0.731 ** | 4.491 | -1.521 *** | 14.028 | -0.410 | 0.758 |
| Wealth quintile: 2 | -0.177 | 0.719 | -1.554 *** | 13.760 | -0.794 ** | 2.753 |
| Wealth quintile: 3 | -0.304 | 0.480 | -1.305 *** | 8.772 | -0.972 ** | 4.714 |
| Wealth quintile: 4 | -0.374 | 1.600 | 0.005 | 10.461 | -0.550 | 0.844 |
| Total household tasks | 0.020 | 1.973 | 0.022 | 2.424 | -0.002 | 0.121 |
| Bank branch in community | 0.056 | 0.170 | 0.304 | 0.386 | 0.850 | 0.910 |
| Market/bazaar in community | -0.008 | 0.426 | -0.180 | 0.346 | -0.296 | 0.719 |
| Kirana market in community | 0.144 | 1.454 | -0.429 | 0.628 | 0.152 | 0.774 |
| Middle school in community | 0.017 | 0.058 | -0.056 | 0.122 | -0.722 | 0.852 |
| Cooperative in community | 0.036 | 0.072 | -0.070 | 0.145 | -0.573 | 0.417 |
| Nearest town < 20 km | -0.042 | 1.508 | -0.159 | 0.072 | -0.568 | 1.923 |
| Block fixed effects | n.a. | 60.506 | n.a. | 15.852 | n.a. | 54.137 |
| N | 1624 |  | 969 |  | 970 |  |

\*\*\* $p < .01$ , \*\* $p < .05$ , \* $p < .1$
<sup>a</sup>Shapley-Owen decomposition of contribution to R-squared. Grouped R-squareds presented for the block dummies.

## Ethical review information

The protocol, informed consent forms, and study questionnaires were approved by the Institutional Review Boards (IRB) of the International Food Policy Research Institute in Washington, DC, USA; the Institute of Health Economics, University of Dhaka, Bangladesh; Centre for Media Studies, Delhi, India; and the National Health Research Council, Kathmandu, Nepal.

